# Development of a Mixture Model (SMM) Allowing for Smoothing Functions of Trajectories

**DOI:** 10.1101/2019.12.13.19014928

**Authors:** Ming Ding, Jorge E. Chavarro, Garrett M. Fitzmaurice

## Abstract

In the health and social sciences, two types of mixture model have been widely used by researchers to identify heterogeneous trajectories of participants within a population: latent class growth analysis (LCGA) and the growth mixture model (GMM). Both methods parametrically model trajectories of individuals, and capture latent trajectory classes, by using an expectation-maximization (E-M) algorithm. However, parametric modeling of trajectories using polynomial functions or monotonic spline functions results in limited flexibility for modelling trajectories; as a result, group membership may not be classified accurately due to model misspecification. In this paper, we propose a mixture model (SMM) allowing for smoothing functions of trajectories using a modified E-M algorithm. In the E step, participants are reassigned to only one group for which the estimated trajectory is the most similar to the observed one; in the M step, trajectories are fitted using generalized additive mixed models (GAMM) with smoothing functions of time. This modified E-M algorithm is straightforward to implement using the recently released “gamm4” macro in R. The SMM can incorporate time-varying covariates and be applied to longitudinal data with normal, Bernoulli, and Poisson distributions. Simulation results show favorable performance of the SMM in terms of classification of group membership. The proposed method is illustrated by its application to body mass index data of individuals followed from adolescence to young adulthood and the relationship with incidence of cardiometabolic disease.

## 1. INTRODUCTION

In the health and social sciences, mixture model has been used to identify heterogeneous trajectories of participants within a population.^1,2^ Two types of parametric mixture models have been developed and widely used by researchers: the growth mixture model (GMM) proposed by Muthen *et al* and latent class growth analysis (LCGA) proposed by Nagin *et al*.^3,4^ Both models allow different sets of parameter values for mixture components corresponding to different unobserved subgroups of individuals, and capture latent trajectory classes with different growth curves by using an expectation-maximization (E-M) algorithm. The difference between the two models lies in that GMM allows for variation across individuals within the same group while LCGA assumes individuals within groups are homogenous.^5^ However, both GMM and LCGA model covariates as polynomial functions, although recent extensions of GMM allow for monotonically non-decreasing I-spline functions ^6^, resulting in limited flexibility for modelling trajectories. Furthermore, group membership may not be classified accurately to represent the true unobserved subgroups due to model misspecification.

Semiparametric mixture models with smoothing functions of covariates have been developed.^7,8^ However, some limitations of these models are that they can only be applied to normally distributed data, and there is no existing software to implement them. Smoothing splines have been widely applied to model covariates nonlinearly in longitudinal data, e.g., in generalized linear models ^9,10^ and generalized estimating equations.^11-14^ To allow for mixed effects in models for longitudinal data, they were further implemented within generalized linear mixed models (GLMM),^15-18^ creating a new class of models referred to as generalized additive mixed models (GAMM).^19^ GAMM provides nonparametric functions of covariates and uses random effects to account for correlation in longitudinal data.^19^ Although the computation of GAMM is highly intensive, it is straightforward to apply using the package ‘gamm4’ in R, which performs well not only for continuous data, but also for binary and count data.^20^ As GAMM can model covariates with high flexibility and accounts for within-individual correlation, a natural extension is to consider a GAMM applied to trajectory analysis. Similar to GMM and LCGA which are essentially a combination of GLMM with latent class analysis, in this paper we develop a smoothing mixture model (SMM) allowing for smoothing functions of trajectories by combining GAMM with latent class analysis. As parameter estimation of our model based on maximum likelihood estimation (MLE) would be very computationally demanding, we use a modified E-M algorithm to simplify parameter estimation: in the E step, rather than assigning to all groups with different membership probabilities, each individual is assigned to only one group with the highest membership probability; in the M step, the existing package ‘gamm4’ is applied directly to estimate model parameters within each group.^20^ By adopting this algorithm, we avoid having to maximize intractable likelihoods, greatly simplifying model development and application.

## 2. METHOD

Similar to the E-M algorithm, the group assignment was achieved by iterating the modified M and E steps. In the first iteration, we assigned participants to a predefined number of groups according to the mean value of a participant’s trajectory. In the M step, we fit GAMM models with smoothing functions of time for all groups and obtained predicted trajectories from all GAMM models for each individual. In the E step, we reassigned participants to the group for which the estimated trajectory was the most similar to the observed one. We iterated the modified E-M algorithm until group membership no longer changes. The details of the method are outlined below.

### 2.1. Notation

Let the sample consists of *n* individuals. Consider the data on individual *i* to consist of a vector ***Y***_***i***_ of p repeated measurements over time ***T***_***i***_, and a (q × p) matrix ***X***_***i***_ of covariates. The components of ***Y***_***i***_ could be continuous, count, or binary data. For example, ***Y***_***i***_ could be repeated measures of lifestyle factors across adulthood. In the 1st iteration, we divided participants into *k* groups according to the mean value of a participant’s trajectory. Let individual *i* be assigned to the *m*th group.

### 2.2. The modified E-M algorithm of group assignment

Step 1. Maximization step (M step)

- Using individuals in the *m*th group, fit a nonparametric GAMM model with smoothing spline of time (e.g., using ‘gamm4’ package in R 3.2.5) 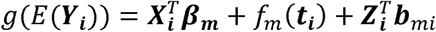, where ***Y***_***i***_ is the outcome for the *i*th individual in group *m, g* (.) is the link function, ***t***_***i***_ is the vectors of times of measurement of ***Y***_***i***_, *f*_*m*_ (.) is a smoothing function of time, ***β***_***m***_ is a q × 1 vector of regression coefficients associated with covariates ***X***_*i*_, ***b***_*mi*_ are independent b × 1 vectors of random effects associated with covariates ***Z***_***i***_(the latter usually a subset of ***X***_***i***_ and ***t***_***i***_).
- Although the vector ***Y***_***i***_ for individual *i* only contributes to the parameter estimation in the *m*th group, we obtain *k* vectors of predicted value ***Ŷ***_***i*(1)**_, ***Ŷ***_***i*(2)**_, …, ***Ŷ***_***i*(*k*)**_ estimated from GAMMs fitted in the *1*st, *2*nd, …, *k*th groups, respectively. Given estimates from the *k* fitted GAMMs, ***Ŷ***_***i*(*k*)**_ can be expressed as 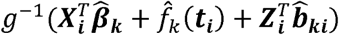 Step 2. Expectation step (E step)
- For individual, we obtain the log likelihood contributions *𝓁*_*i*(1)_, *𝓁*_*i*(2)_, …, *𝓁*_*i*(*k*)_ of individual *i*’s trajectory of responses belonging to the *1*st, *2*nd, …, *k*th groups. It can be shown for continuous, binary, and count data that *𝓁*_*i*(*k*)_ ≈ - (***Y***_***i***_ − ***Ŷ***_***i*(*k*)**_)^′^*Wi*(***Y***_***i***_ − ***Ŷ***_***i*(*k*)**_), where *W*_*i*_ = diag{*α* (ø)*v*{***Ŷ***_***i*(*k*)**_,}}^−1^, a p × p diagonal matrix. — Formula (1)

**Derivation of formula (1):** For the *k*th group, the *𝓁*_*i*(*k*)_conditional on *b*_*ki*_ can be expressed as 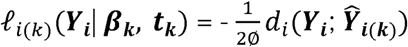, where *d*_*i*_(***Y***_***i***_; ***Ŷ***_***i*(*k*)**_) is the deviance. The deviance statistic ***d***_***i***_(***Y***_***i***_; ***Ŷ***_***i*(*k*)**_)can be approximated by the Pearson chi-square statistic

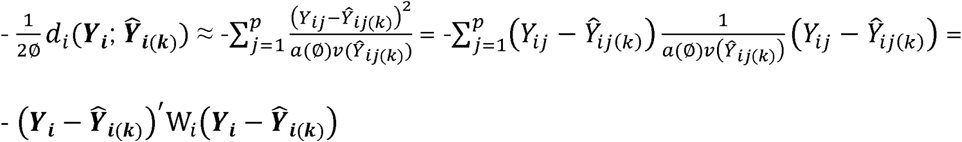 ***Ŷ***_***i*(1)**_ can be estimated directly from the fitted GAMM in the *k*th group. For exponential family distributions, ø is the dispersion parameter, *a* (ø) is a function of the dispersion parameter, and *v* (.) is the variance function. Specifically, if *Y*_*ij*_ belongs to normal distribution N (µ, *δ*^2^), ø is *δ*^2^, *a*(ø) is *δ*^2^, and *v* (.) is 1. If *Y*_*ij*_ belongs to Bernoulli distribution B (1, p), ø is 1, *a*(*δ*) is 1, and *v* (.) is p(1-p). If *Y*_*ij*_ belongs to Poisson distribution P(µ), ø is 1, *a*(*δ*) is 1, and the *v* (.) is µ.
- We compare *𝓁*_*i*(1)_, *𝓁*_*i*(2)_, …, *𝓁*_*i*(*k*)_. In a departure from the traditional algorithm where in the E step individual *i* is reassigned to all of the *k* groups with different probabilities, in our proposal, individual *i* is reassigned to the group with the largest likelihood, *𝓁*_*i*_.= max (*𝓁*_*i*(*1*)_, *𝓁*_*i*(*k*)_,*…* ,*𝓁*_*i*(*k*)_). Step 3. Iteration of E-M steps. Iterate the modified E-M steps until model converges. Model convergence is determined when the group membership for all individuals no longer change, and the sum of the largest likelihood for all individuals 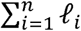 remains the same.

### 2.3. Number of groups

The Bayesian Information Criterion (BIC) is used to compare model fit assuming different numbers of groups.^21-23^ Consider that we divide all individuals’ trajectories into *k* groups. Upon model convergence, the log *L*_1_, log *L*_2_, …, log *L*_*k*_ are the log likelihoods estimated using GAMM in the *1*st, *2*nd, …, *k* th groups, and *p*_1_, *p*_2_, …, *p*_*k*_ are the respective degrees of freedom. The BIC for our mixture model is defined as

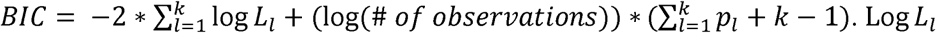

and *p*_*l*_ can be obtained using the MER and EDF commands from the ‘gamm4’ output in R 3.2.5, which apply to data with normal, Bernoulli, and Poisson distributions.

## 3. SIMULATION

We conducted a simulation study to assess the performance of the proposed SMM in terms of classification of group membership.

### 3.1. Simulation strategy

We simulated datasets with 2, 3, and 4 groups of trajectories, with 100 individuals in each group and 20 observations for each individual at time points evenly spaced between 0 to 1. We simulated each individual’s trajectory using functions shown below, which could produce a variety of trajectory shapes with high flexibility and be used to generate data with normal, Bernoulli, and Poisson distributions. Specifically, for individual *i* at time *j* in the *m*th group, *g*(*mean* (*Y*_*mij*_)) was generated from 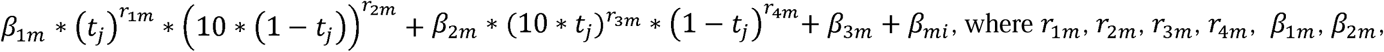 and *β*_3*m*_ were group-specific parameter, and *β*_*mi*_ was an individual-specific parameter that belonged to a normal distribution with mean 0 and standard deviation 1. For data from a normal distribution, *g* (.) was identity function, and we included a random error *ε*_*mij*_ ∼ N(0, 1) to (*Y*_*mij*_) to simulate *Y*_*mij*_. For Bernoulli distribution, *g* (.) was logit function. We obtained 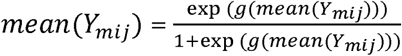, and sampled each observation *Y*_*mij*_ from a Bernoulli distribution with *mean* (*Y*_*mij*_) For Poisson distribution, g (.) was log function. We obtained *mean* (*Y*_*mij*_) = exp (*g*(*mean* (*Y*_*mij*_))), and sampled each observation *Y*_*mij*_ from a Poisson distribution with *mean* (*Y*_*mij*_). For normal, Bernoulli, and Poisson distributions, we generated trajectories with high, medium, and low separation by assigning different values to the standard deviation of *β*_*mi*_. Compared to trajectories with high separation, trajectories with low separation had larger individual-specific random-effects. The parameter setting with high, medium, and low separation is shown in **Table S1** for data with normal, Bernoulli, and Poisson distributions. The mean trajectories simulated are shown in **Figures 1-3** for data with normal, Bernoulli, and Poisson distributions.

**Figure 1.**
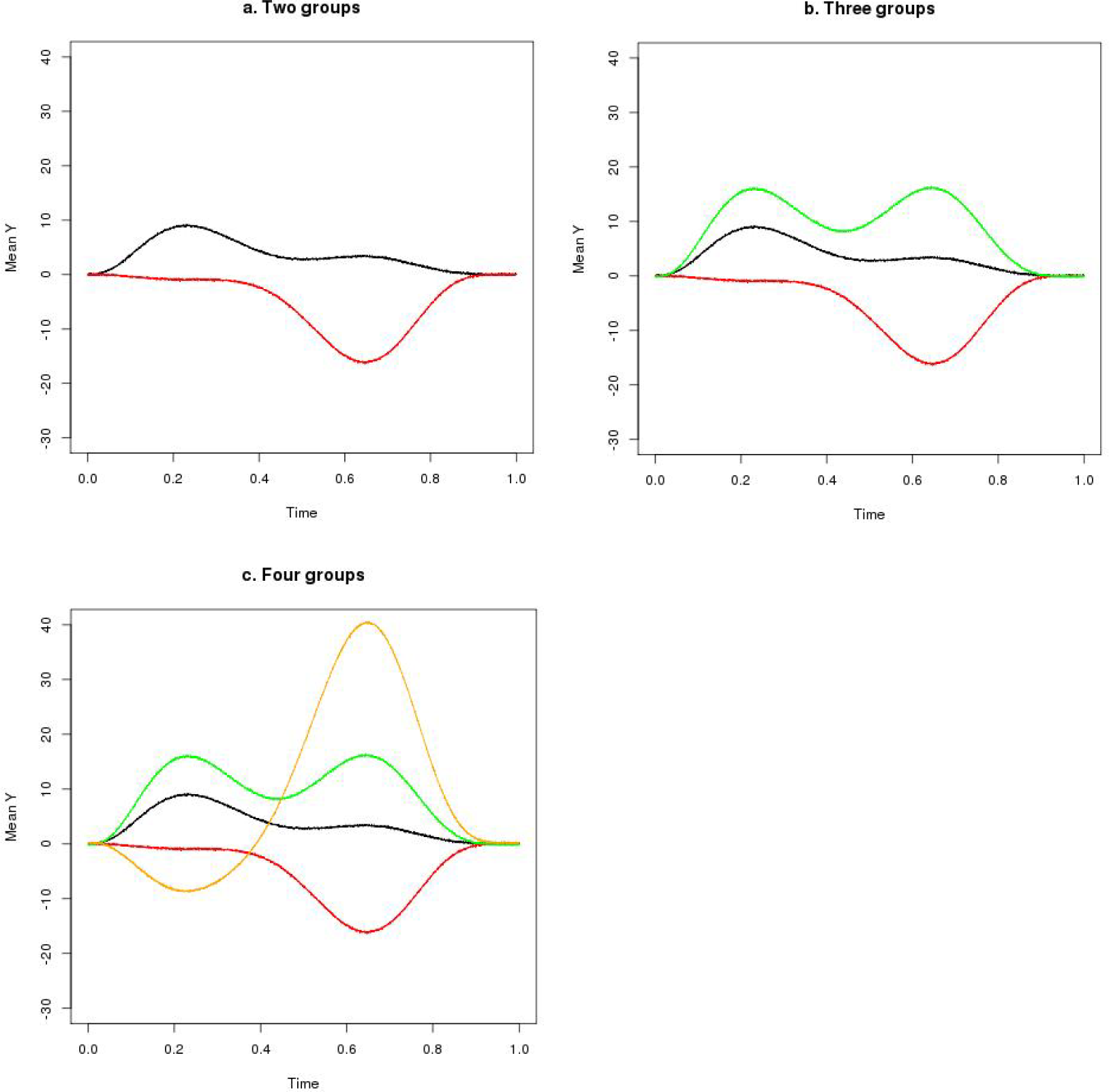
Simulated longitudinal data with normal distribution.

**Figure 2.**
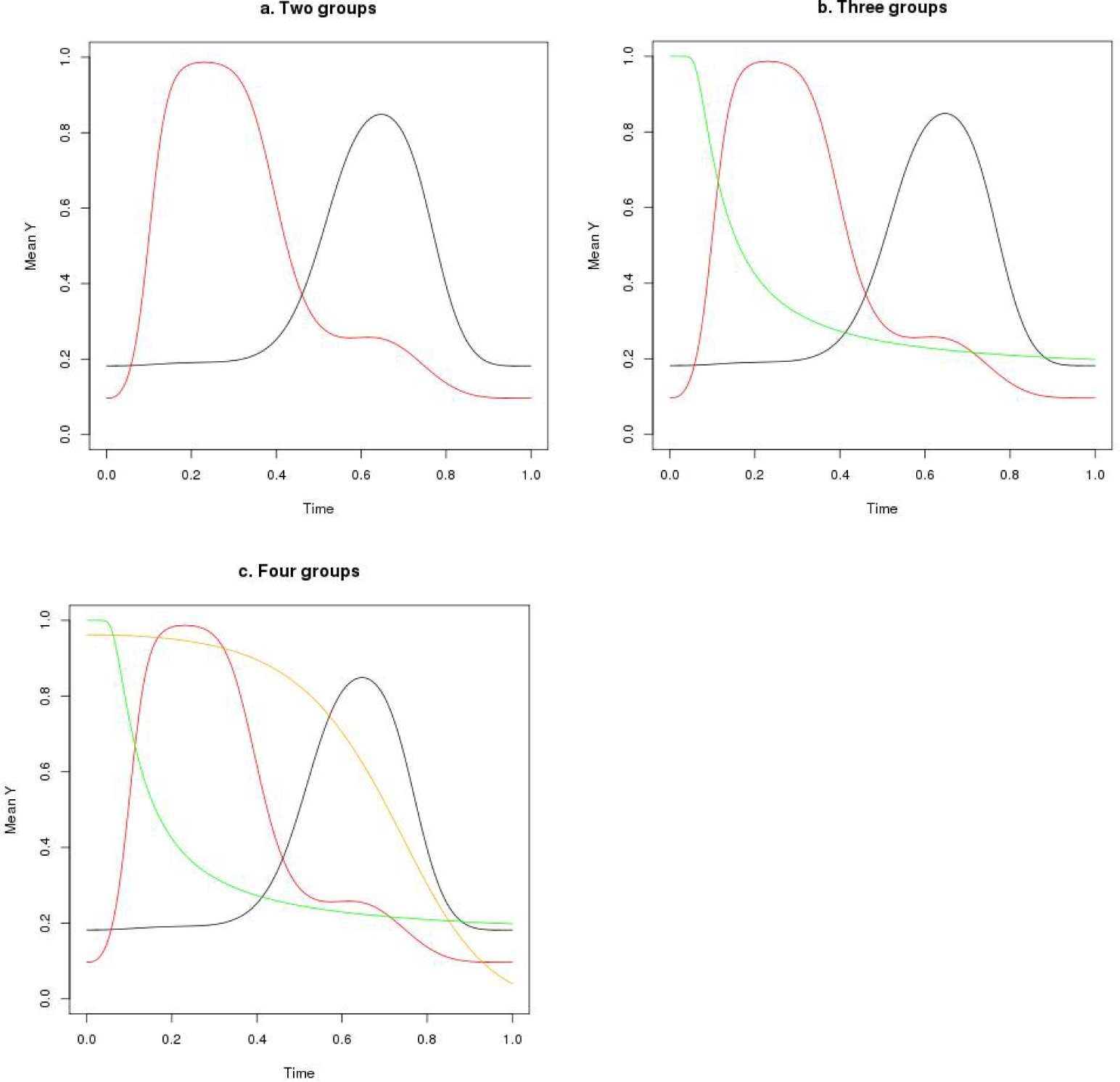
Simulated longitudinal data with Bernoulli distribution (we presented data simulated with median separation).

**Figure 3.**
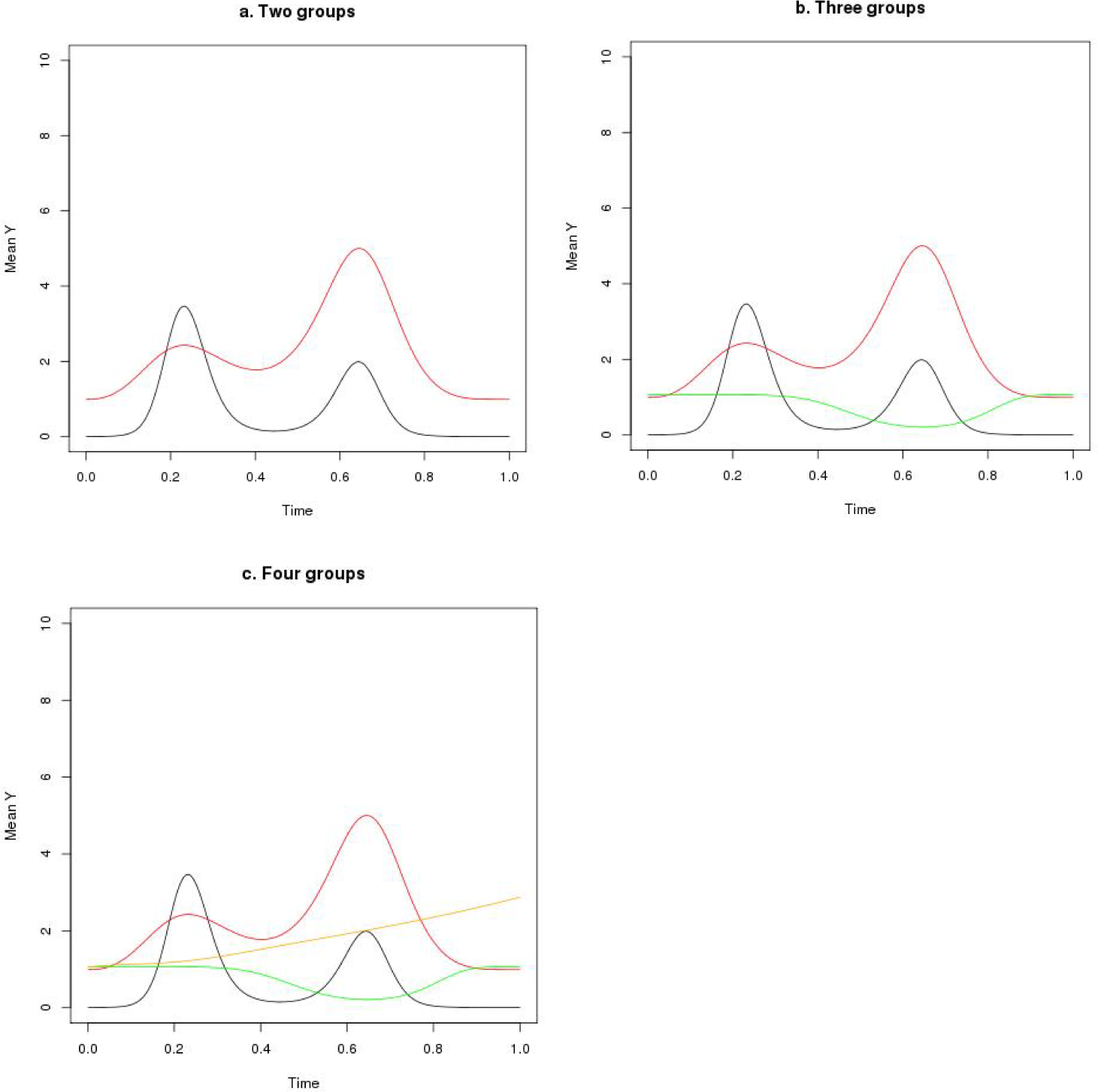
Simulated longitudinal data with Poisson distribution (we presented data simulated with median separation).

### 3.2 Model fit

We have developed an R script for the proposed smoothing mixture model, and the code is shown in supplemental material. We initially assigned individuals to different groups based on the rank of average value of Y across time points. To obtain model estimates, the modified E-M algorithm was iterated for 20 times, which suggested model convergence as indicated by BIC from each iteration. We randomly simulated the datasets with normal, Bernoulli, and Poisson distributions for 1000 times and fit the SMM to each of the simulated datasets. We obtained the percentage of individuals that were correctly classified (individuals that remained in the same group), the correlation coefficient between predicted and observed values (Pearson correlation for data with normal distribution and Spearman correlation for data with Bernoulli and Poisson distributions), and the number of groups identified as indicated by the lowest BIC (for simulated data with two trajectory groups, we compared BIC assuming one, two and three groups; for simulated data with three trajectory groups, we compared BIC assuming two, three, and four groups; for data with four trajectory groups, we compared BIC assuming three, four, and five groups).

### 3.3. Results

We simulated trajectories with high flexibility to evaluate the performance of SMM in modeling non-linear trajectories. We further simulated trajectory groups with high, medium, and low separation to assess the performance of our model in classifying group membership and identifying number of groups. As to data with normal distribution, our model successfully classified most of the individuals in settings with high and medium separation between groups (**Table 1**). Even for low separation, with a large standard deviation of 5 for individual-specific random-effects, our model successfully classified 70-80% of individuals. Consistently, we found strong correlation between observed and predicted values, and the predicted trajectories were highly similar to the simulated trajectories (**Figure S1**). Our model identified correct number of groups in the scenario of high separation, however, it tended to identify more groups with medium and low separation. As to data with Bernoulli distribution, our model assigned group membership with high accuracy for trajectories with high separation (**Table 2**), and the predicted trajectories were similar to simulated ones with high separation (**Figure S2**) although the correlation between observed and predicted values was not particularly strong. In addition, our model tended to identify more groups of trajectories in scenario of medium and low separation. For data with Poisson distribution, the SMM correctly assigned most of the individuals in datasets with high and medium separation (**Table 3**). Our model fitted the data well as indicated by the high similarity between predicted and simulated trajectories; however, the correlation between observed and predicted values was not particularly strong (**Figure S3**). Similar to data with normal and Bernoulli distribution, our model identified more groups in scenarios of medium and low separation.

**Table 1.** Model fit for longitudinal data with normal distribution simulated for 1000 times using smoothing mixture model (SMM).

| | Random effect, $\beta_{mi}$ | Percentage of individuals correctly classified (% , median) | Pearson correlation coefficient between predicted and observed value (mean) | Number of groups identified as indicated by lowest BIC (percentage of datasets over 1000 simulated datasets) * |
| --- | --- | --- | --- | --- |
| **Two groups** |  |  |  |  |
| High separation | N(0, 0.5) | 100 | 0.98 | 2 (100%) |
| Medium separation | N(0, 2) | 100 | 0.94 | 2 (82%), 3(18%) |
| Low separation | N(0, 5) | 86 | 0.82 | 2 (5%), 3 (95%) |
| **Three groups** |  |  |  |  |
| High separation | N(0, 0.5) | 100 | 0.99 | 3 (100%) |
| Medium separation | N(0, 2) | 98 | 0.96 | 3 (35%), 4 (65%) |
| Low separation | N(0, 5) | 75 | 0.89 | 3 (15%), 4 (85%) |
| **Four groups** |  |  |  |  |
| High separation | N(0, 0.5) | 100 | 0.99 | 4 (100%) |
| Medium separation | N(0, 2) | 99 | 0.97 | 3 (10%), 4 (53%), 5 (37%) |
| Low separation | N(0, 5) | 81 | 0.93 | 4 (38%), 5 (62%) |

\*BIC: Bayesian information criterion.

**Table 2.** Model fit for longitudinal data with Bernoulli distribution simulated for 1000 times using smoothing mixture model (SMM).

| | Random effect, $\beta_{mi}$ | Percentage of individuals correctly classified (% , median) | Spearman correlation coefficient between predicted and observed value (mean) | Number of groups identified as indicated by lowest BIC (percentage of datasets over 1000 simulated datasets) * |
| --- | --- | --- | --- | --- |
| **Two groups** |  |  |  |  |
| High separation | N(0, 0.5) | 100 | 0.65 | 2 (77%), 3 (23%) |
| Medium separation | N(0, 2) | 65 | 0.53 | 2 (8%), 3 (92%) |
| Low separation | N(0, 4) | 53 | 0.64 | 3 (100%) |
| **Three groups** |  |  |  |  |
| High separation | N(0, 0.5) | 99 | 0.61 | 3 (69%), 4 (31%) |
| Medium separation | N(0, 2) | 60 | 0.60 | 4 (100%) |
| Low separation | N(0, 4) | 50 | 0.69 | 3 (1%), 4(99%) |
| **Four groups** |  |  |  |  |
| High separation | N(0, 0.5) | 97 | 0.70 | 4 (63%), 5 (37%) |
| Medium separation | N(0, 2) | 56 | 0.67 | 5 (100%) |
| Low separation | N(0, 4) | 44 | 0.73 | 5(100%) |

\*BIC: Bayesian information criterion.

**Table 3.** Model fit for longitudinal data with Poisson distribution simulated for 1000 times using smoothing mixture model (SMM).

| | Random effect, $\beta_{mi}$ | Percentage of individuals correctly classified (% , median) | Spearman correlation coefficient between predicted and observed value (mean) | Number of groups identified as indicated by lowest BIC (percentage of datasets over 1000 simulated datasets) * |
| --- | --- | --- | --- | --- |
| **Two groups** |  |  |  |  |
| High separation | N(0, 0.1) | 100 | 0.75 | 2 (38%), 3 (62%) |
| Medium separation | N(0, 0.3) | 94 | 0.73 | 2 (3%), 3 (97%) |
| Low separation | N(0, 0.5) | 81 | 0.70 | 2 (15%), 3 (85%) |
| **Three groups** |  |  |  |  |
| High separation | N(0, 0.1) | 97 | 0.68 | 3 (59%), 4 (41%) |
| Medium separation | N(0, 0.3) | 90 | 0.65 | 3 (10%), 4 (90%) |
| Low separation | N(0, 0.5) | 73 | 0.59 | 3 (5%), 4 (95%) |
| **Four groups** |  |  |  |  |
| High separation | N(0, 0.1) | 96 | 0.65 | 4 (12%), 5 (88%) |
| Medium separation | N(0, 0.3) | 85 | 0.63 | 4 (15%), 5 (85%) |
| Low separation | N(0, 0.5) | 66 | 0.62 | 4 (7%), 5 (93%) |

\*BIC: Bayesian information criterion.

## 4. APPLICATIONS

The Growing-up Today Study (GUTS) was established in 1996 when women participating in the Nurses’ Health Study II (NHSII) were invited to enroll their children aged 9 to 14 years into this new cohort. A total of 16,882 children responded to the baseline questionnaires. Participants have been followed up with yearly self-administered follow-up questionnaires between 1997 and 2001 and with biennial questionnaires thereafter through 2013. GUTS participants reported their height and weight at baseline, and updated these data on follow-up questionnaires. Adolescents have been found to be able to provide valid reports of height and weight ^24-26^. Body mass index (BMI) was calculated as the ratio of weight (kg) to height (m) squared. In adolescents (<18 y), obesity was defined as a BMI at or above the age- and sex-specific cutoffs proposed by the International Obesity Task Force (IOTF) ^24^. In adults (≥18 y), obesity was defined as BMI ≥30 kg/m^2^. In 2010 and 2013, participants were asked to report in questionnaire whether they developed diabetes, hypertension, and hypercholesterolemia, and the year of diagnosis (<1996, 1996-1999, 2000-2005, and 2006-2013). We created a composite outcome cardiometabolic incidence, defined as incidence of diabetes, hypertension, or hypercholesterolemia. To minimize reverse causation, we excluded participants who were diagnosed before 1999 and censored BMI reported after diagnosis of cardiometabolic disease. We further excluded individuals with less than two BMI measurements. For participants who were siblings, we randomly chose one participant to avoid between-person correlation of BMI. In total, we included 10,743 participants among whom 1043 cases of cardiometabolic disease. We collected information on total energy intake and physical activity in 1996, 1997, 1998, and 2001 using self-reported questionnaire. We identified BMI trajectories of GUTS participants using LCGA, GMM, and SMM. We assumed random effects for SMM, and iterated the E-M algorithm for 100 times, which suggested model convergence as indicated by BIC from each iteration. The LCGA was fitted using the “traj” command in STATA (Version 11, StataCorp, TX),^27^ and we modeled trajectories with cubic polynomials using LCGA. The GMM was fitted using the “LCMM” package in R 3.5.0,^6^ and we modeled the trajectories using I-splines with 5 knots at quantiles. We compared the SMM to the LCGA and GMM in terms of log likelihood (LLK), BIC, and trajectories delineated using the models. We examined associations of identified trajectories with risk of cardiometabolic disease using logistic regression.

We delineated trajectories of BMI from 9 to 30 years adjusting for time-stable (sex) and time-varying covariates (total energy intake and physical activity), and SMM showed the highest LLK and lowest BIC in comparison to LCGA and GMM (**Table 4**). Although BIC decreased with increase in number of groups using all three models, we chose three groups to select a parsimonious model that captures a high amount of information. Of the three trajectories identified using SMM, one had consistently high BMI through the follow-up period (on average, participants in this group were obese at 47% times of assessment); one had consistently medium BMI (participants in this group were obese at 3% times of assessment); and the other trajectory had consistently low BMI (participants in this group were not obese at any time of assessment) (**Figure 4**). For all three trajectories, BMI increased with age; and the growth rate of BMI was high in adolescence and slowed after adulthood (age ≥18 years). In general, the trajectories estimated using LCGA were similar to those estimated using SMM, however, the trajectory of the consistently high BMI group was not as smooth as that produced using SMM. For trajectories generated using GMM, although BMI increased with age, the increase rate of BMI accelerated with age and this trend persisted even after adulthood. We examined associations of trajectories identified using the three methods with risk of cardiometabolic diseases, and found that the odds ratio of cardiometablic diseases was significantly higher in medium and high BMI groups in a dose-response manner comparing to the consistently low BMI group (**Table 5**).

**Table 4.** Comparison of model fit for trajectories of body mass index (BMI) using latent class growth analysis (LCGA), growth mixture model (GMM), and smoothing mixture model (SMM) in the Growing-up Today Study (GUTS).

| Number of<br>groups classified | Model | Log likelihood | BIC |
| --- | --- | --- | --- |
| 2 | LCGA | -153622 | 307364 |
| 2 | GMM | -149440 | 299048 |
| 2 | SMM | -129152 | 258565 |
| 3 | LCGA | -145499 | 291183 |
| 3 | GMM | -142274 | 284780 |
| 3 | SMM | -124450 | 249284 |
| 4 | LCGA | -141649 | 283548 |
| 4 | GMM | -139180 | 278657 |
| 4 | SMM | -121378 | 243262 |
| 5 | LCGA | -139645 | 279605 |
| 5 | GMM | -137662 | 275687 |
| 5 | SMM | -119257 | 239145 |

Trajectories adjusted for sex and time-varying variables including total energy intake and physical activity.

**Table 5.**
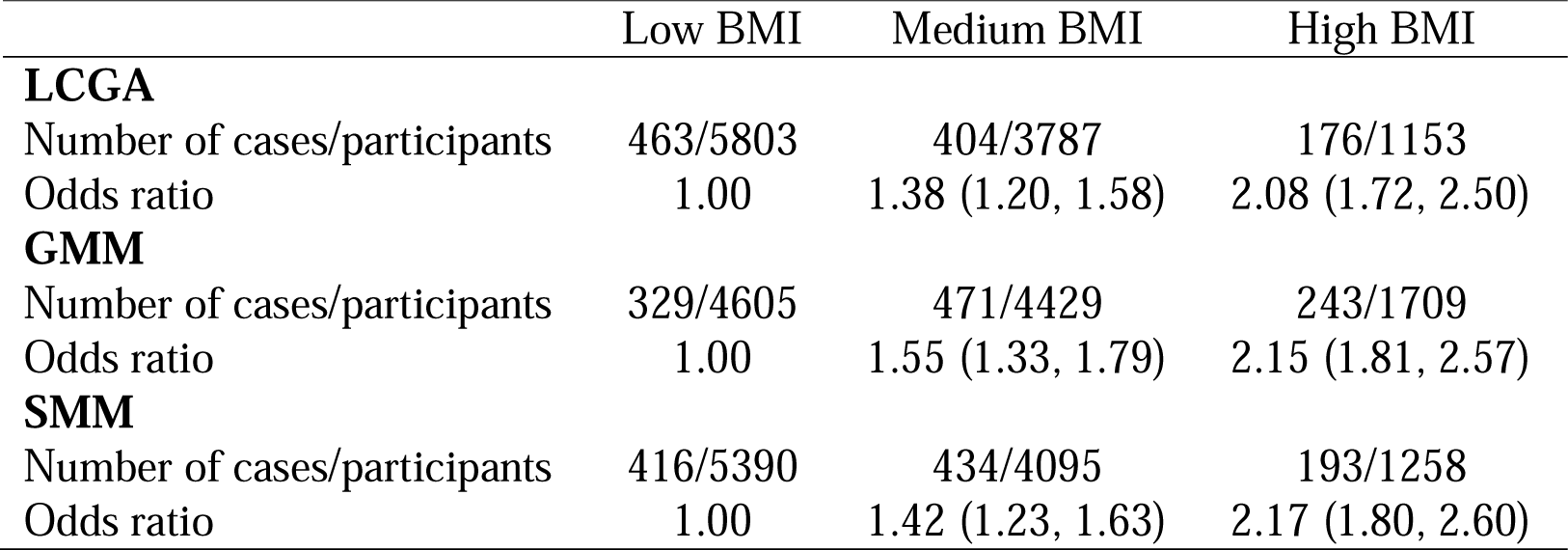
Associations of trajectories of body mass index with risk of cardiometabolic disease using latent class growth analysis (LCGA), growth mixture model (GMM), and smoothing mixture model (SMM) in the Growing-up Today Study (GUTS).

**Figure 4.**
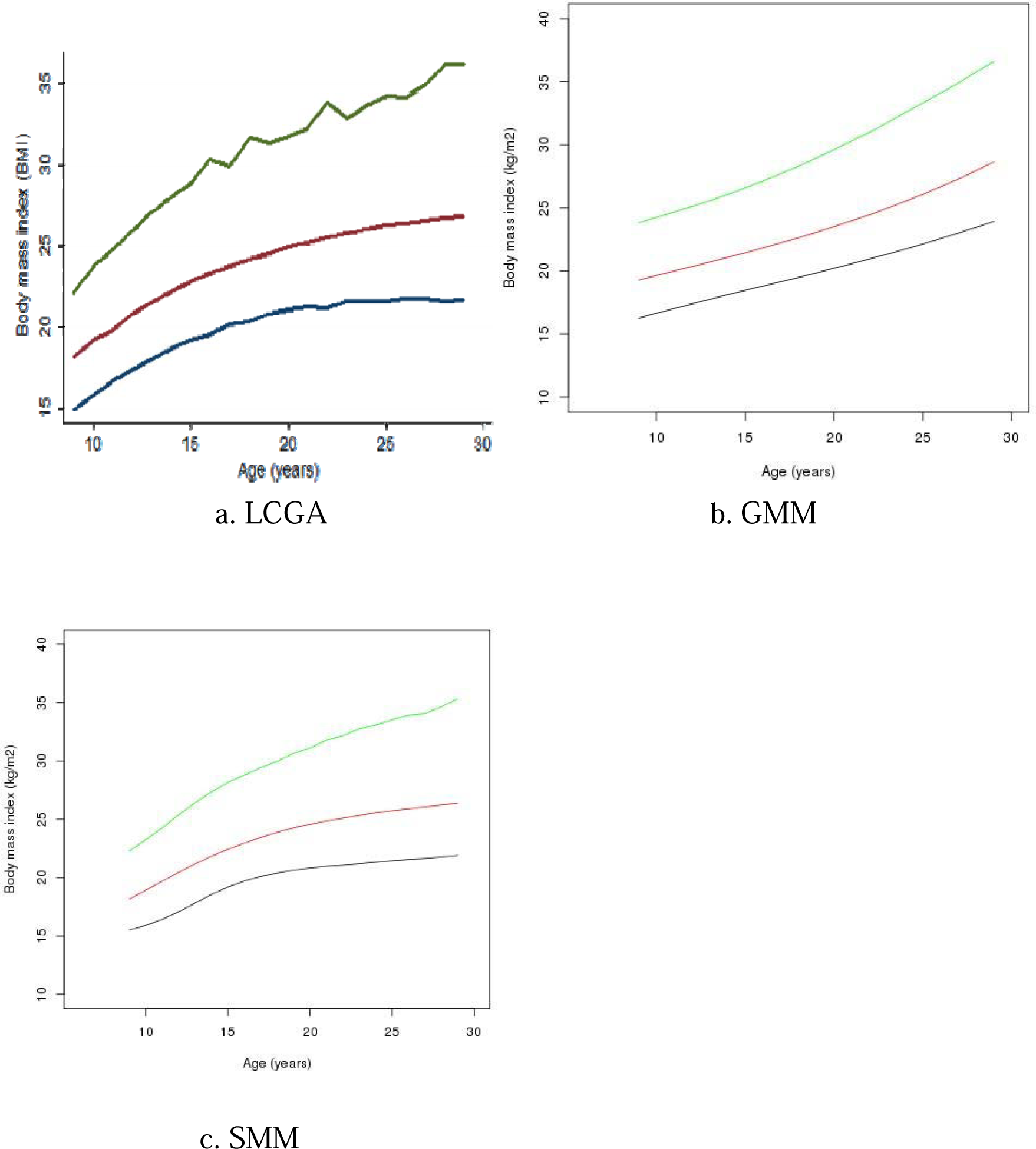
Trajectories of body mass index delineated using latent class growth analysis (LCGA), growth mixture model (GMM), and smoothing mixture model (SMM) in the Growing-up Today Study (GUTS).

## 5. DISCUSSION

In this study, we developed a mixture model allowing for smoothing functions of trajectories using a modified E-M algorithm. In the E step, we reassigned participants to only one group for which the estimated trajectory was the most similar to the observed one, and in the M step, we utilized the recently released “gamm4” macro to fit GAMM models with smoothing functions of time. Comparing to existing mixture models including LCGA and GMM, the advantages of SMM lie in modeling trajectories with high flexibility and the application to data with normal, Bernoulli, and Poisson distributions.

In a simulation study, we simulated highly flexible longitudinal data with low, medium, and high separations to evaluate performance of our model in classifying group membership, delineating trajectories, and identifying number of groups. As to data with a normal distribution and medium to high separation, our model achieved outstanding performance in assigning group membership and delineating trajectories, which indicates potentially wide application of our model to non-linear longitudinal trends with normal distributed data. As to data with Bernoulli and Poisson, our model performed reasonably well in assigning group membership and estimating trajectories when there was medium to high separation. This is a distinct advantage of our model in comparison to other mixture models. Instead of adopting logit or log link function and using generalized estimating equations for model estimation, GMM treats data with Bernoulli and Poisson distributions as ordinal outcomes and uses threshold functions for model estimation.^6^ As GMM assumes a threshold effect between time and observed outcome, the trajectories estimated are less flexible. In contrast, when our model is applied to data with Bernoulli and Poisson distributions it takes advantage of the most recently published “gamm4” macro to fit GAMM. Compared to the earlier “gamm” macro, gamm4 does not use penalized quasi-likelihood (PQL) for model estimation, and thus gives better performance to binary and low mean count data.^20^

While recognizing our model has some potential advantages for highly flexible modelling and convenient application to data with different distributions, we acknowledge several limitations of the SMM. First, using BIC, our model tended to identify too many groups, even in the scenario of medium separation that indicates relatively low heterogeneity in trajectories. However, this is also a common issue for mixture models, which occurred for LCGA and GMM in the application section of our paper. Nagin *et al* holds the viewpoint that the most basic test of adequacy is whether the final model adequately addresses the research question,^1^ and Bauer *et al* suggests that the number and shape of groups should be guided by a priori expectation.^28^ Second, the application of our method can be computationally demanding, given that in the M step, the “gamm4” macro needs to estimate between-person random effects; moreover, the modified E-M algorithm can require a large number of iterations. Thus, for our method, computation time may be a concern for datasets with large sample size and multiple repeated measurements. Third, by using an iterative, hill-climbing procedure, the modified E-M algorithm may converge to a local maximum, and initial assignment of group membership affects the local maximum identified and the speed of convergence. In our study, we used rank of mean value of individual’s trajectory to assign initial group membership. Comparing to other initialization procedures such as random starting values and k-means algorithm, this method is simple to apply and avoids multiple initial assignment to reduce computation time.^29^

In the application, we identified trajectories of BMI across adolescence and young adulthood using SMM in comparison to LCGA and GMM. The BMI trajectories identified using SMM increased with age, and the growth rate of BMI slowed down after individual entering adulthood. This is consistent with the depictions of growth charts of U.S. adolescence, which shows that growth spurts begin at 10-12 years, last throughout adolescence, and end at 18-20 years with the cessation of rapid growth ^30^. Our study adds new knowledge that individuals with high, medium, and low BMI at baseline share similar growth patterns, and individuals with high BMI in adolescence are highly likely to remain obese in young adulthood. Moreover, we found that individuals with high BMI in adolescence are associated with higher risk of cardiometabolic diseases in early adulthood. The potential mechanism is that childhood obesity is associated with chronic inflammation and elevation of inflammation biomarkers ^31^. This can lead to insulin resistance, dyslipidemia, and high blood pressure, which enhance the development and progression of cardiometabolic diseases. Overall, the application study showed the importance of adopting a healthy BMI in adolescence and maintaining a low-BMI growth trajectory in prevention of obesity and cardiometabolic diseases in young adulthood. By applying our model to life-course epidemiology, our model has significant public health implication in enhancing our understanding of how early exposure over time affects health outcomes. Given the high flexibility of trajectory modeling and the convenient application to data with different distributions, we expect our model has broad application in health and social sciences.

In the application of our model to trajectories of BMI across adolescence and young adulthood, we would like to clarify two concerns. First, in the M step of the modified E-M algorithm, instead of estimating parameters using an overall likelihood that fitted the whole data, we split the dataset into several groups and fitted GAMM within each group. Thus, for covariates included in the SMM, the underlying assumption is that associations of covariates with BMI might differ across trajectory groups. Second, we censored BMI after development of cardiometabolic disease. This might lead to different patterns of exposure distribution associated with outcome, as individuals who did not develop disease would have exposures repeatedly measured at more time points. Thus, how to extend our method to deal with time-to-event outcome would be a highly interesting direction for future research.

In summary, we developed a mixture model allowing for smoothing functions of trajectories using a modified E-M algorithm. Our model can be applied to normal, Bernoulli, and Poisson distributed data, and has favorable performance in terms of classification of group membership. Application of our model to life course epidemiology shows its potentially wide application in health and social sciences.

## Data Availability

The data is available upon request.

## 6. SUPPLEMENTARY MATERIALS

R script of our smoothing mixture model is also provided in the online Supplementary Materials.

## ACKNOWLEDGEMENTS

This work was supported by grants R03 AG060247, P30-DK046200 and U01-HL145386 from the National Institutes of Health. We appreciate Dr. Xihong Lin’s support in application of grant R03 AG060247.

## DATA SHARING

The data that support the findings of this study are available on request from the corresponding author. The data are not publicly available due to privacy or ethical restrictions.

## Reference

1. Nagin DS, Odgers CL. Group-based trajectory modeling in clinical research. Annual review of clinical psychology 2010;6:109–38.

2. Marie R, Monique S, Kieron O. The Evolution of the Study of Life Trajectories in Social Sciences over the Past Five Years: A State of the Art Review. Advances in Mental Health 2010;9:190–210.

3. Nagin DS, Tremblay RE. Analyzing developmental trajectories of distinct but related behaviors: a group-based method. Psychological methods 2001;6:18–34.

4. Muthen B, Shedden K. Finite mixture modeling with mixture outcomes using the EM algorithm. Biometrics 1999;55:463–9.

5. Berlin KS, Parra GR, Williams NA. An introduction to latent variable mixture modeling (part 2): longitudinal latent class growth analysis and growth mixture models. Journal of pediatric psychology 2014;39:188–203.

6. Proust-Lima C, Philipps V, Liquet B. Estimation of Extended Mixed Models Using Latent Classes and Latent Processes: The R Package lcmm.. Journal of Statistical Software 2017;78:1–56.

7. Huang Y, Qiu H, Yan C. Semiparametric Mixture Modeling for Skewed Longitudinal Data: A Bayesian Approach. Ann Biom Biostat 2015;2:1011.

8. Nummi T, Salonen J, Koskinen L, Pan J. A semiparametric mixture regression model for longitudinal data. Journal of Statistical Theory and Practice 2017.

9. Hastie T, Tibshirani R. Generalized Additive Models. Statistical Science 1986;1:297–318.

10. Nelder JA, Wedderburn RWM. Generalized Linear Models. Journal of the Royal Statistical Society Series A (General) 1972;135:370–84.

11. Rice JA, Silverman BW. Estimating the Mean and Covariance Structure Nonparametrically when the Data are Curves. Journal of the Royal Statistical Society Series B (Methodological) 1991;53:233–43.

12. Liang KY, Zeger SL. Longitudinal Data Analysis Using Generalized Linear Models. Biometrika 1986;73:13–22.

13. Wild CJ, Yee TW. Additive Extensions to Generalized Estimation Equation Methods. Journal of the Royal Statistical Society Series B (Methodological) 1996;58:711–25.

14. Berhane K, Tibshirani R. Generalized additive models for longitudinal data. The Canadian Journal of Statistics 1998;26:517–35.

15. Breslow NE, Clayton DG. Approximate Inference in Generalized Linear Mixed Models. Journal of the American Statistical Association 1993;88:9–25.

16. Zeger SL, Diggle PJ. Semiparametric models for longitudinal data with application to CD4 cell numbers in HIV seroconverters. Biometrics 1994;50:689–99.

17. Zhang D, Lin X, Raz J, Sowers MF. Semiparametric Stochastic Mixed Models for Longitudinal Data. Journal of the American Statistical Association 1998;93:710–9.

18. Verbyla AP, Cullis BR, Kenward MG, Welham SJ. The Analysis of Designed Experiments and Longitudinal Data by Using Smoothing Splines. Journal of the Royal Statistical Society Series C (Applied Statistics) 1999;48:269–311.

19. Lin X, Zhang D. Inference in generalized additive mixed models by using smoothing splines. Journal of the Royal Statistical Society Series B (Statistical Methodology) 1999;61:381–400.

20. Wood S, Scheipl F. Package ‘gamm4’. https://cran.r-project.org/web/packages/gamm4/gamm4.pdf. 2016.

21. Schwarz G. Estimating the Dimension of a Model. The Annals of Statistics 1978;6:461–4.

22. Brame R, Nagin DS, Wasserman L. Exploring Some Analytical Characteristics of Finite Mixture Models. Journal of Quantitative Criminology 2006;22:31–59.

23. Nylund KL, Asparouhov T, Muthén BO. Deciding on the Number of Classes in Latent Class Analysis and Growth Mixture Modeling: A Monte Carlo Simulation Study. Structural Equation Modeling 2007;14:535–69.

24. Goodman E, Hinden BR, Khandelwal S. Accuracy of teen and parental reports of obesity and body mass index. Pediatrics 2000;106:52–8.

25. Strauss RS. Comparison of measured and self-reported weight and height in a cross-sectional sample of young adolescents. International journal of obesity and related metabolic disorders : journal of the International Association for the Study of Obesity 1999;23:904–8.

26. Field AE, Aneja P, Rosner B. The validity of self-reported weight change among adolescents and young adults. Obesity (Silver Spring) 2007;15:2357–64.

27. Jones BL, Nagin DS. A Note on a Stata Plugin for Estimating Group-based Trajectory Models. Sociological Methods & Research 2013;42:1–6.

28. Bauer DJ. Observations on the use of growth mixture models in psychological research. Multivariate behavioral research 2007;42:757–86.

29. Shireman E, Steinley D, Brusco MJ. Examining the effect of initialization strategies on the performance of Gaussian mixture modeling. Behavior research methods 2017;49:282–93.

30. Kuczmarski RJ, Ogden CL, Guo SS, et al. 2000 CDC Growth Charts for the United States: methods and development. Vital and health statistics Series 11, Data from the National Health Survey 2002:1–190.

31. Weihrauch-Bluher S, Schwarz P, Klusmann JH. Childhood obesity: increased risk for cardiometabolic disease and cancer in adulthood. Metabolism: clinical and experimental 2019;92:147–52.

